# Biomarkers in the prediction of multimorbidity: scoping review

**DOI:** 10.1101/2020.11.25.20214999

**Authors:** EA Spencer, GA Ford, MS Chan, R Perera, CJ Heneghan

## Abstract

**Background:** Multimorbidity presents an increasing challenge in the global ageing population. Predicting its development is necessary to help design and deliver effective healthcare.

**Objective:** This scoping review aimed to collate and present the body of published evidence on biomarkers and multimorbidity, identifying what work has been done and what gaps remain.

**Methods:** We searched the electronic databases MEDLINE, Register of Controlled Trials (Cochrane CENTRAL), CINAHL, PsycINFO, EMBASE, Scopus, Web of Science and TRIP database up until 11 August 2020 and hand-searched the reference lists of included articles.

**Results:** We found 34 relevant studies including 12 reporting prospective data and 22 reporting cross-sectional data. These encompassed 14 studies on serum biomarkers, 2 on molecular biomarkers, 7 on physiological biomarkers, 8 on body size biomarkers and 3 on brain function biomarkers. Most studies were undertaken in European or North American populations. There was a broadly consistent finding that obesity was associated with increased multimorbidity. Other results were more varied, reflecting the diverse range of biomarkers investigated, and lack of standardisation of multimorbidity outcome definitions.

Longitudinal studies have been set up that are maturing and further evidence can be expected over time.

**Conclusion:** There has been limited research on biomarkers to predict the development of multimorbidity, with minimal investigation of putative biomarkers identified in basic research. High quality research studies in this area are needed to progress the development of targeted interventions to prevent or delay the onset of multimorbidity.

## Background

Multimorbidity is the co-occurrence of two or more diseases in the same individual where each must be a non-communicable disease (NCD), or a mental health disorder, or a long duration infectious disease. [1] NCDs account for a significant amount of morbidity and mortality: 71% of all deaths globally and 15 million premature deaths. [2]

Multimorbidity is more common with ageing, affecting two-thirds of the elderly. Around one in four UK adults have two or more long-term conditions, often described as multimorbidity, and this rises to two-thirds of people aged 65 years or over. [3] Evidence shows that multimorbidity decreases quality of life and increases morbidity and mortality. [4][5]

Early recognition, screening and treatment is needed to minimise the risks to individuals, while reducing costs to the healthcare system. In those with long-term conditions, UK health and social care expenditure is estimated to take up around £7 in every £10 of the total spend. [6] Globally, multimorbidity also leads to high levels of out-of-pocket expenditure for patients. [7]

One way to identify individuals at an earlier stage is through the use of biological and physiological markers that predict various combinations of multimorbidity. [8] The accumulation of chronic disease at older ages has been associated with various markers that may act as an early warning sign to better target interventions, aid identification of preventive strategies and reduce the associated burden of multimorbidity through better treatment.

We undertook a scoping review of the literature to map the literature on biomarkers in multimorbidity, identify key concepts and gaps in the research literature; assess the current levels of evidence and the quality of evidence available and to describe what the evidence shows on the relationship of biomarkers or physiological markers and incident multimorbidity in adults.

## Methods

We published the protocol and any necessary amendments [9], and wrote the report according to the PRISMA statement on reporting scoping reviews. [10]

### Eligibility criteria

We included studies with adults with two or more long term conditions compared to those without multimorbidity (i.e. those with no long term conditions or only one long term condition). For a study to be eligible, study participants had to be aged 18 years and older; the measurement of the biomarker (the exposure measure) had to predate the outcome of multimorbidity and had to be measured in adulthood.

We included cohort studies (prospective and retrospective); RCTs from which we were able to extract data from the control arm; and case control studies. We also included systematic reviews which contain studies meeting our eligibility criteria not otherwise identified in our searches; and where applicable, guidelines or reports from the grey literature. We also included protocols or records of relevant ongoing trials and prospective studies. We excluded cross-sectional studies; case series and case reports.

We included studies that use a biomarker, defined as ‘measurable and quantifiable biological parameters (e.g., specific enzyme concentration, specific hormone concentration, specific gene phenotype distribution in a population, presence of biological substances) which serve as indices for health- and physiology-related assessments, such as disease risk, psychiatric disorders.’ (see the PubMed MESH database for the full list) [11] We also included studies using physiological markers defined as measurable and quantifiable physiological parameters (e.g., oral temperature, blood pressure (BP), heart rate, body weight etc.). Thus, studies of physical activity per se were not included, but studies of physical function, such as handgrip or walking speed, were eligible.

We excluded biomarkers directly related to the presence or absence of any health outcome (for example prior disease, medication and treatment), health behaviours, socioeconomic factors, environmental factors, sex and chronological age.

We excluded studies that did not specify the marker or do not have an objective measure of the marker, or that related the marker to only one specific disease. We excluded studies not in English.

Due to the paucity of studies on biomarkers and incident multimorbidity found, we also included studies reporting data on the cross-sectional relationship between biomarkers and multimorbidity.

### Information sources

We searched the following electronic databases: MEDLINE, Register of Controlled Trials (Cochrane CENTRAL), CINAHL, PsycINFO, EMBASE, Scopus, Web of Science, TRIP database. The search was conducted from the beginning of the databases up until the search date, 11 August 2020. We also screened the reference lists of included studies for possible additional studies to assess for eligibility.

The literature search was based on the following search terms, adapted to each database (full details are given in Web Appendix 1. Literature search strategy): multimorbidity, comorbidity, multi comorbidity, multiple diseases, multiple morbidities, multiple pathology, disease clustering, chronic diseases, severity of illness illness; biomarker, physiological markers, markers, biological, molecular markers, genetics, molecular, biochemical markers.

### Selection of sources of evidence

Two reviewers (ES, CH) independently screened the initial retrieved studies for eligibility. The first screen selected based on study title; the next screen reviewed title and abstract; the next screen used full texts. Inclusion and exclusion of retrieved studies are shown in a flow diagram, Figure 1.

**Figure 1.**
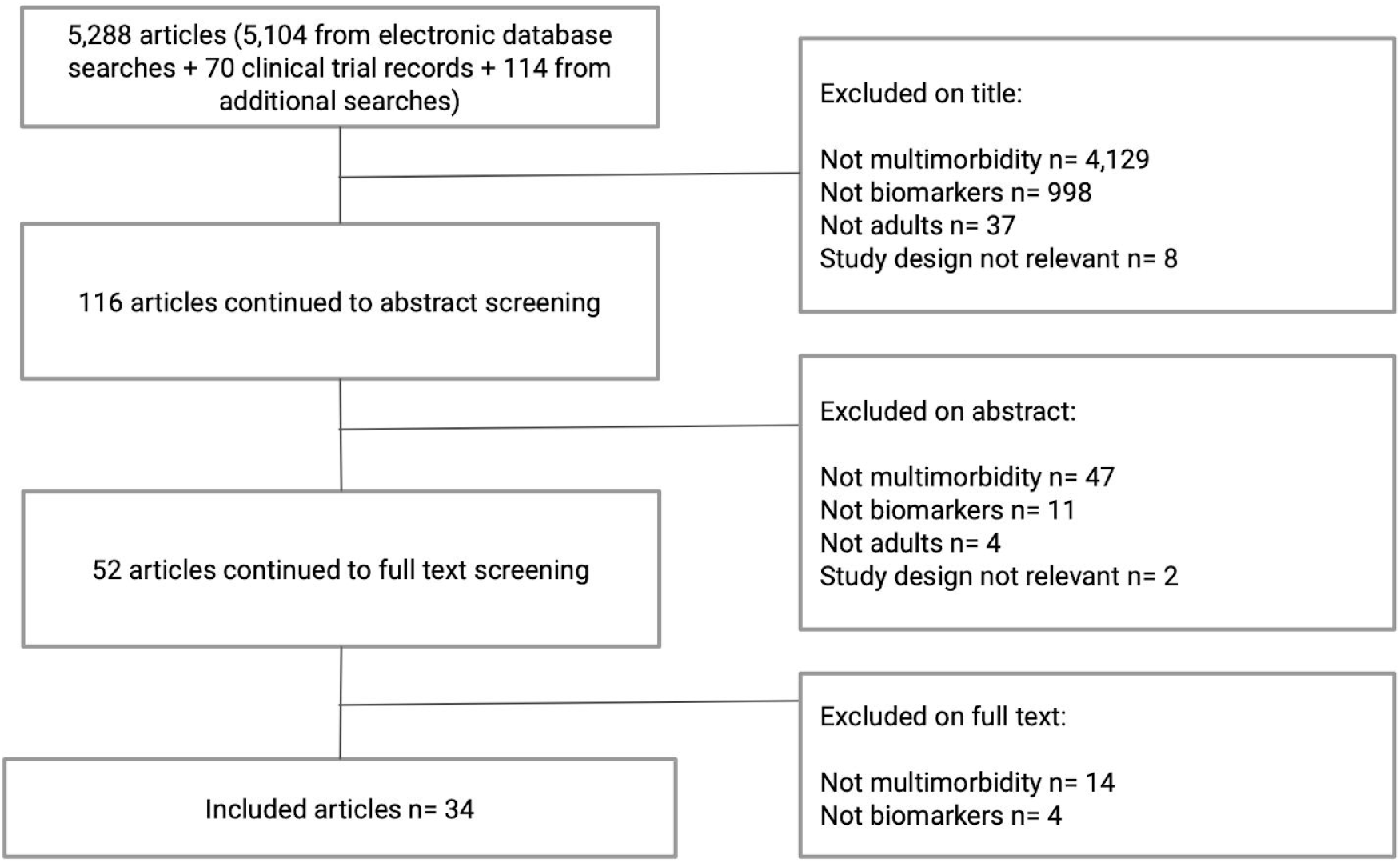
Flow chart of studies in the literature search

### Data extraction and mapping

We assessed the level of evidence using the OCEBM levels of evidence tool We produced a table of characteristics by biomarker grouping. We extracted data on the type of study, country and setting, publication year, sample size, baseline characteristics of the population: age, sex, diseases at baseline and multimorbidity outcomes at follow up, with ascertainment and measurement methods. We planned to use the ICD-11 classification [12] to group rarely reported conditions and to group closely associated conditions where needed, but this was not necessary. We report the relationship of exposure to the outcome as reported in the original paper. These data were extracted by one reviewer (ES) and checked by a second reviewer (CH).

## Results

We screened 5,288 records and of these included 34 eligible studies; see Figure 1 flow chart.

Sample size ranged from 58 (Kahl 2005) to 223,286 (Wong 2014) participants. The 34 included studies were done in seventeen countries. Most were done in the USA, Italy and the UK, and two studies took place across different countries: Stirland 2019 was a European, prospective cohort study done at 12 sites across Europe, and Kivimaki 2017 was a pooled analysis of data from 120,813 participants in the USA and Europe.

**Table 1** shows the level of evidence found for this scoping review: we found no systematic review of randomized trials, nor any randomized trials. Twelve prospective studies (n= 377,626 participants) met our inclusion criteria with data on biomarkers and multimorbidity as an outcome; we also found 22 cross-sectional studies (n= 282,827 participants). Three prospective studies also reported cross-sectional results on multimorbidity (Fabbri 2015a, Fabbri 2015b, Niedzwiedz 2019).

**Table 1.** Level of evidence of included studies.

| OCEBM evidence category | This scoping review: biomarkers in multimorbidity |
| --- | --- |
| Research question: Is this early detection test worthwhile? | What is the available evidence on biological and physiological markers and incident multimorbidity in adults? |
| Systematic review of randomized trials | none identified |
| Randomized trial | none identified |
| Non-randomized controlled cohort/follow-up study | n= 12 included prospective studies |
| Case-series, case-control, or historically controlled studies | n= 22 included cross-sectional studies |
| Mechanism-based reasoning | excluded from this report |

**Table 2** reports the characteristics of included studies, by category of biomarker.

### (i) Serum biomarkers

Three prospective studies reported on serum biomarkers and multimorbidity (Fabbri 2015a, Issa 2020 and Perez 2019).

Fabbri and colleagues examined inflammatory and hormonal biomarkers in an elderly group in Italy and showed that higher baseline IL-6, and steeper increase of IL-6 levels, were significantly and independently associated with multimorbidity over the 9 year follow-up (Fabbri 2015a).

Issa and colleagues investigated incident cardiovascular comorbidities among patients in Denmark with newly diagnosed or longstanding rheumatoid arthritis, and measured serum and synovial fluid MFAP4 at study inception. During a follow-up of four years, MFAP4 correlated positively with stroke events, systolic blood pressure and levels of HDL cholesterol. In multivariate analysis, adjusting for age, sex and smoking, only systolic blood pressure remained significantly associated with MFAP4.

Perez investigated serum glutathione in an elderly group in Sweden, and over a follow-up of 6 years, found lower levels of baseline total serum glutathione were associated with a higher rate of multimorbidity development (Perez 2019).

We identified 11 cross-sectional studies looking at serum biomarkers and multimorbidity, done in Australia, Europe, India, Singapore and South Korea. Three were among outpatients undergoing medical treatment, six were an analysis of an ongoing prospective cohort, and one study was of inpatients in a psychiatric treatment centre. The biomarkers ranged from bone mineral density to inflammatory and nutritional markers.

Some of these cross-sectional studies used a definition of multimorbidity related to the conditions of focus in the study; for example, in a group of breast cancer patients, multimorbidity was a composite of fatigue, arthralgia and insomnia; in a study of patients with COPD, patients with diabetes mellitus in addition to COPD were compared with patients with COPD only (Hyun 2019); in the study of psychiatric patients, those with borderline personality disorder and major depressive disorder were compared with those with borderline personality disorder alone (Kahl 2005); in a small study of patients with TB, those with concurrent diabetes mellitus were compared with those with TB alone (Kumar 2019). These studies, therefore, focussed more on comorbidity than on the concept of multimorbidity per se.

In contrast, some of these cross-sectional studies set out to examine multiple morbidities and used a comprehensive list of possible chronic conditions to create a multimorbidity score, sometimes using a standard index, e.g. Cervellati et al used a standard multimorbidity score incorporating possible items across a number of body systems, the CIRS-CI index (Cervellati 2015). Fabbri et al used a comprehensive multimorbidity score (Fabbri 2015a); Martin-Ruiz and coworkers also used a comprehensive multimorbidity disease count (Martin-Ruiz 2011).

A small study of breast cancer patients found that inflammatory biomarkers in serum (including C-reactive protein, vitamin D-binding protein) were strongly associated with multimorbidity categorised as a composite variable of arthralgia, insomnia and fatigue concurrent with breast cancer (Bauml 2015). An Italian study on outpatients with cognitive decline reported that serum levels of hydroperoxides were positively correlated with multimorbidity in controls and in the mild cognitive impairment group but not in late onset Alzheimer disease patients (Cervellati 2015).

Cross sectional data from the ongoing Italian prospective study by Fabbri and colleagues, looking at inflammatory and hormonal biomarkers, showed that higher serum IL-6, IL-1ra, TNF-α receptor II, and lower DHEA were associated cross-sectionally with higher number of diseases, independent of age, sex, body mass index, and education (Fabbri 2015a). Another study among community elderly in Italy examined the relationships of serum C-reactive protein, lipoprotein (a) and cystatin C levels and number of diseases present; it reported that higher values of all the three biomarkers correlated with a higher number of chronic conditions, both when dichotomized as high versus normal and as continuous variables (Garrafa 2017).

A study of elderly men in Australia showed that having low serum 25-hydroxyvitamin D was associated with greater likelihood of having 4 or more chronic conditions compared with zero to 3 conditions (Hirani 2014). Moo and coworkers found that among nearly 800 mostly elderly patients admitted to hospital for a low impact hip fracture, there was no correlation between serum vitamin D level and age-adjusted Charlson Comorbidity Index (Moo 2020).

A study of outpatients being treated for respiratory disease in Seoul, South Korea, reported that among people with COPD, patients with high plasma fibrinogen concentrations and normal 25-OH vitamin D levels had a significantly higher incidence of diabetes mellitus than did the other patients (Hyun 2019).

A small study of German inpatients (mean age 26 years) with borderline personality disorder diagnosis looked at bone mineral density and markers of bone turnover in relation to the presence or absence of major depressive disorder concurrent with borderline personality disorder; bone mineral density was significantly lower in the group with borderline personality disorder and comorbid major depressive disorder than in the group with borderline personality disorder alone. Values of crosslaps, osteocalcin, serum cortisol, TNF-α, and interleukin-6 were significantly higher in the patients with borderline disorder plus current major depressive episode than in the healthy subjects; patients with borderline personality disorder who did not have current or lifetime depression displayed no alterations of either bone mineral density or the immunological and hormonal measures examined (Kahl 2005).

A small study of patients with TB in Chennai, India (n=132) showed that TB-with diabetes was associated with elevated systemic levels of circulating monocyte activation markers compared with TB alone (Kumar 2019).

A UK study of elderly participants aged 85 years or older investigated the cross-sectional associations at baseline of 74 candidate biomarkers and morbidities: many biomarkers were associated with disease; the 10 most strongly associated were: N-terminal proB-type natriuretic peptide, handgrip strength, blood pressure (BP), the timed up-and-go test, FEV, haematocrit, haemoglobin, red blood cell count, free T3, and vitamin D (Martin-Ruiz 2011). A cohort in the Netherlands assessed plasma 25-hydroxyvitamin D3 and found cross-sectional dose-response relationships of lower vitamin D levels and increased morbidity (Meems 2015). A study of middle aged to elderly aged adults in Germany tested metabolic, inflammatory and oxidative stress markers in serum, including HbA1c, C-reactive protein, cholesterol, and derivatives of reactive oxygen metabolites. The results showed that all the blood biomarkers except thiol were positively associated with multimorbidity (Schöttker 2016).

### (ii) Molecular biomarkers

We found two studies reporting the relationships between molecular biomarkers and incident multimorbidity, both prospective. Lee and coauthors (2009) reported no association between ACE I/D genotype and multimorbidity (COPD with asthma/IHD/hypertension vs COPD alone). Niedzwiedz 2019 investigated telomere length (telomeres are the protective ends on chromosomes; they shorten over time and have been hypothesised to affect the rate of ageing); in their data, telomere length at baseline was not associated with the development of multimorbidity over 2 to 6 years of follow-up. Cross-sectional data on molecular biomarkers from a prospective study by Niedzwiedz et al showed some evidence that telomere length was related to reduced likelihood of multimorbidity, but these associations were not statistically significant using a binary multimorbidity variable as the outcome (Niedzwiedz 2019).

### (iii) Physiological biomarkers

Strandberg (2014) reported results from a Finnish cohort study of men, in whom blood pressure was measured at baseline and morbidities tracked over 48 years of follow-up: baseline blood pressure had a graded and highly significant association with the number of comorbidities that developed over time.

We found six cross-sectional studies reporting on physiological biomarkers and multimorbidity. Two indicated that low handgrip strength was related to higher multimorbidity; the relationship was stronger among men than women (Amaral 2015, Cheung 2013).

Ishizaki et al reported cross-sectional results from an ongoing longitudinal study, and in just over 2.5 thousand participants aged 60 years and older, handgrip strength was associated with multiple chronic diseases and multimorbidity in men and women, after adjustment for confounding factors. There was a linear trend of association with the number of chronic diseases in men, but not in women (Ishizaki 2020).

Cross-sectional findings on blood pressure and multimorbidity were mixed. In a UK based study, lower blood pressure was associated with higher multimorbidity (Sarkar 2015). In a small study of patients with panic disorder, blood pressure and heart rate were higher in the multimorbid group (diagnosed with both panic disorder and major depression) compared with those with either panic disorder alone or major depression alone (Townsend 1998). In a large study of Hong Kong residents, there was a strong relationship of poor blood pressure control with increasing multimorbidity (Wong 2014).

### (iv) Body size biomarkers

We identified nine studies assessing body size and multimorbidity. Six prospective studies reported the relationship between BMI and incident multimorbidity (Booth 2014, Fabbri 2015b, Humphreys 2018, Kivimaki 2017, Mounce 2018, Xu 2019). These studies were based in Europe, Australia and the USA. In all six studies, higher BMI at baseline was associated with greater risk of multimorbidity. Xu 2019 reported that weight gain was associated with increased risk of developing multimorbidity.

We found three cross-sectional studies of body size biomarkers and multimorbidity: a US survey of middle aged and older adults showed the odds of multimorbidity were increased as the classification and reported duration of overweight/obesity worsened (Dankel 2018); a study of older adults in Italy showed that obesity was associated with higher morbidity count, but not in overweight compared with normal weight participants (Fabbri 2015b); and a study of a relatively young population in Pakistan reported a significant linear trend of increasing morbidities with increasing body weight categories (Khan 2017).

### (v) Brain function biomarkers

Three cross-sectional studies investigated biomarkers of brain function and multimorbidity and reported inconsistent results. A small study of elderly participants in France showed that accumulation of multimorbidity was associated with neuroimaging markers of Alzheimer disease neurodegeneration, but not with amyloid deposition (Mendes 2018). A study of cerebrospinal fluid amyloid-42 among adults without dementia found that people with multimorbidity were approximately 40% less likely to be amyloid positive than people with one or no conditions (Stirland 2019). A study of cognitively normal elderly USA residents found that multimorbidity were associated with abnormal Alzheimer disease signature meta-region of interest 18F-FDG hypometabolism, and with abnormal Alzheimer disease signature MRI cortical thickness, but was not associated with amyloid accumulation (Vassilaki 2016).

#### Research implications

Table 3 shows the research priorities as reported within the included papers. Most recommendations made non-specific statements that more research in this area is necessary to establish associations and to begin to understand the biological processes involved in multimorbidity. The published prospective data on this topic is limited and existing prospective studies with biological repositories could look at incident multimorbidity and how that relates to biological markers. Even the cross-sectional data are limited, with the exception perhaps of body size as a biomarker, where a number of studies have demonstrated a relationship between overweight and increased multimorbidity.

**Table 3.** Research implications.

| Study ID | Research implications, as reported in the included article. |
| --- | --- |
| **Prospective cohort studies** |  |
| Booth 2014 | no research recommendations |
| Fabbri 2015b | "...further investigations in larger, more-diverse populations in terms of culture and body composition characteristics (including underweight or extremely obese individuals) are required to confirm the generalizability of the findings..." |
| Humphreys 2018 | no research recommendations |
| Issa 2019 | "Further studies are needed to elucidate the potential of MFAP4 as a risk indicator for cardiovascular morbidity in RA" |
| Kivimaki 2017 | no research recommendations |
| Lee 2009 | no research recommendations |
| Mounce 2018 | "Further research on multimorbidity clusters is needed to clarify relevant mechanisms: co-prevalence with increasing age (eg, osteoporosis and arthritis) vs shared pathophysiology (eg, hypertension and coronary artery disease). Future work should also account for morbidity burden - for example, the level of control of such conditions as diabetes, ischemic heart disease, and arthritis - which may provide a more meaningful measure of morbidity to clinicians and patients than using counts of conditions." |
| Niedzwiedz 2019 | "Further prospective studies with larger samples and studies that assess the direction of causality (e.g. via Mendelian Randomisation) are needed to investigate the relationship between telomere length and specific types of multimorbidity, such as psychiatric disorders and cardiovascular disease." |
| Perez 2019 | "...further investigation is needed to better understand the causal relationship between tGSH levels, biological changes, aging, and the development of multimorbidity." |
| Strandberg 2014 | no research recommendations |
| Xu 2019 | "...more prospective studies are warranted to understand which combination of chronic conditions together most frequently, and how the conditions combination progress over time among women with body weight gain or loss." |
| **Cross-sectional studies** |  |
| Amaral 2015 | "The study also indicates the need for further epidemiological research to improve our understanding of the findings based on clinical parameters for diseases and focused on specific age groups, explaining the differences between men and women and contributing to proposals of reference values and cutoff points for health risks." |
| Cervellati 2015 | no research recommendations |
| Cheung 2013 | "Since it [handgrip strength] is an objective parameter assessing muscle strength, which is affecting and controlled by multiple physiological system, future clinical trial is warranted to investigate its usefulness in intervention or prevention of multimorbidity, and hence possibly preventing premature mortality." |
| Dankel 2018 | no research recommendations |
| Fabbri 2015a | no research recommendations |
| Fabbri 2015b | see above |
| Garrafa 2017 | no research recommendations |
| Hyun 2019 | "Additional prospective studies are needed to determine whether vitamin D deficiency leads to the decline in lung function and exacerbation or to determine whether severe COPD provokes vitamin D deficiency." |
| Ishizaki 2019 | "Health assessments that evaluate subjective health status and objective physical performance may provide a better understanding of the complex health status patterns of older adults with multimorbidity and inform the development of health and healthcare management strategies." |
| Kumar 2019 | no research recommendations |
| Meems 2015 | no research recommendations |
| Mendes 2018 | "A future longitudinal analysis in multimorbidity may help to understand the progression of neurodegeneration and amyloid deposition along with possible causality associations." |
| Moo 2020 | no research recommendations |
| Niedzwiedz 2019 | see above |
| Schöttker 2016 | "Further, ideally longitudinal studies are required to corroborate these findings by simultaneous assessment of metabolic, inflammatory, and oxidative stress markers in older adults." |
| Stirland 2019 | "...this result needs further exploration...further research could explore the mechanisms explaining the inverse association between multimorbidity and amyloid positivity." |
| Vassilaki 2016 | "...longitudinal follow-up of the cohort will enable us to identify the effects of multimorbidity on changes in imaging biomarkers." |
| Wong 2014 | "Future research should monitor the trend of multimorbidity among patients with various chronic diseases, and evaluate interventions which are effective to address the complexity of multimorbidity in terms of better disease control." |

Fabbri 2015a pointed out that diverse populations are needed to better understand the generalizability of the results on body size and multimorbidity. Mounce 2018 highlighted the need to distinguish biological mechanisms of multimorbidity from shared pathophysiologies. Several authors noted the difficulty of investigating multimorbidity given the lack of consensus on categories and definitions. Several authors stated the importance of longitudinal data in establishing evidence for causality.

## Discussion

This scoping review shows that currently there is little research published on using biomarkers as early indicators of developing multimorbidity. Among the studies shown here, several were not established with multimorbidity as the focus. Multimorbidity is emerging as an important concept for identifying patient’s needs and treatment and care options, and may also be useful for population health planning and interventions. Therefore it seems necessary for the research community to develop clear questions, consistent definitions and set up high quality prospective studies and ultimately trials to establish how biomarkers can best be used in these contexts.

Regarding the literature we identified, research on serum biomarkers as early indicators of risk of developing multimorbidity seems to be at a relatively early stage; candidate biomarkers are still being sought and some prospective studies set up to look at this relationship are awaiting follow-up to provide results. We found only two studies of molecular risk factors and multimorbidity; this seems an under-studied area.

Among the studies presented here, the most consistent evidence was for the association with BMI: all nine (six prospective) studies showed that higher BMI at baseline was associated with greater risk of multimorbidity. The relatively large number of studies on body size and multimorbidity (incident or cross-sectionally observed) does not necessarily reflect clinical interest in this area, particularly since BMI may be substantially impacted by a range of diseases. These studies may however reflect the relative ease of measuring and correlating body size markers with outcomes.

Results for blood pressure were mixed, and some studies investigated measures of frailty such as low handgrip strength which showed a stronger association with multimorbidity among men than among women. Finally, three varied cross-sectional studies investigated brain function markers, reporting inconsistent results.

Some of the cross-sectional data reported are from ongoing cohort studies; therefore we can anticipate publications of longitudinal data as these studies mature.

One potential barrier in this research area is the lack of standardisation and definitions for multimorbidity. Ermogenous and coauthors discuss multimorbidity in the context of ageing and suggest that consensus is growing that multimorbidity patterns generally fall into three main clusters: cardiometabolic, neuropsychiatric, and musculoskeletal. [15] Using this approach, the cardiovascular cluster might for example comprise coronary heart disease, diabetes, and obesity. A neuropsychiatric cluster could mean an individual has chronic pain, and anxiety and depression. The musculoskeletal cluster could include back pain and osteoporosis. Individuals with multimorbidities in these clusters may also be suffering a range of other conditions. [15]

Another limiting factor is that in order to avoid complexity in understanding intervention effects, clinical trials have often excluded participants with multiple chronic conditions. [16] [17] This reduces applicability to real world clinical settings, and as ageing and multimorbidity increase, it will become essential for clinical trials to include participants with multiple long-term conditions.

Turning the question the other way around, it may be possible to establish biomarkers for healthy ageing, as proposed by Lara and colleagues. [18] Once again, a consistent approach across research studies looking to gain prospective data on incident multimorbidity will be needed.

## Strengths and limitations

We attempted in this scoping review to assess the amount and nature of published evidence on biomarkers and multimorbidity. Ensuring completeness was challenging, given the lack of standardised definitions and approaches in multimorbidity research. Currently, at least 35 multimorbidity indices are available, beyond disease counts, with different components and outcomes. [15] Studies found here did not necessarily describe their work as multimorbidity related. Our search may have missed relevant publications. While the mesh term ‘comorbidity’ has been used since 1990, ‘multimorbidity’ was only introduced as a MESH term in January 2018 [15,16]; pre-2018 studies may not have used the term.

We did not assess quality beyond the level of evidence assessment, as there is little to be gained beyond recognising that cross-sectional studies provide low-quality evidence for inferring causal associations. Prospective studies may be hampered by insufficient sample sizes, non-standardised methods, multiple investigations increasing the type I error rate, and confounding; the studies presented here may suffer from such limitations.

## Recommendations

Establishing clear reporting standards for consensus on categories and definitions of comorbidities would aid research in this area. The establishment of a biomarker registry containing linked evidence with the development of reporting standards could improve the quality of research in this field. Finally, understanding the mechanisms underpinning the biomarker selection will be vital to planning further studies and for establishing the plausibility of causal associations. [17]

## Conclusion

There is currently a limited amount of research on biomarkers and multimorbidity. There is a lack of consensus in outcome criteria, and identification of useful biological and physiological markers is at an early stage.

Many included studies shown here did not set out to examine relationships between biomarkers and multimorbidity. Given the emerging need to comprehend and treat individuals with multiple morbidities, there is a requirement for research that focuses on identifying risk and new ways to target and treat multimorbidity. Additional longitudinal prospective studies focused on biomarkers and multimorbidity are warranted, with consensus on reporting standards and definitions required to aid inference and interpretation of the emerging data.

## Supporting information

Table 2. Characteristics of included studies.

## Data Availability

This is a review of publicly available data published in the scientific literature.

## Acknowledgments

We thank Nia Roberts for her help with the literature searches.

## References

1 Multimorbidity: Technical Series on Safer Primary Care. Geneva: World Health Organization; 2016. Licence: CC BY-NC-SA 3.0 IGO.

2 Non communicable diseases. https://www.who.int/news-room/fact-sheets/detail/noncommunicable-diseases (accessed 15 Nov 2019).

3 England NHS. NHS England " Multimorbidity – the biggest clinical challenge facing the NHS? https://www.england.nhs.uk/blog/dawn-moody-david-bramley/ (accessed 5 Nov 2019).

4 Nunes BP, Flores TR, Mielke GI, Thumé E, Facchini LA. Multimorbidity and mortality in older adults: A systematic review and meta-analysis. Arch Gerontol Geriatr 2016;67:130–8.

5 van den Akker M. MTSSZMSS. Multimorbidity and quality of life: Systematic literature review and meta-analysis. Ageing Research Reviews 2019;53:100903.

6 Long-term conditions and multi-morbidity. The King’s Fund. https://www.kingsfund.org.uk/projects/time-think-differently/trends-disease-and-disability-long-term-conditions-multi-morbidity (accessed 15 Nov 2019).

7 Sum G, Hone T, Atun R, et al. Multimorbidity and out-of-pocket expenditure on medicines: a systematic review. BMJ Glob Health 2018;3:e000505.

8 Ferreira GD, Simões JA, Senaratna C, et al. Physiological markers and multimorbidity. Journal of Comorbidity. 2018;8:2235042X1880698. doi:10.1177/2235042x18806986

9 Evidence-Based Multimorbidity - CEBM. CEBM. 2020. https://www.cebm.net/2020/01/evidence-based-multimorbidity/ (accessed 28 Feb 2020).

10 Tricco AC, Lillie E, Zarin W, et al. PRISMA Extension for Scoping Reviews (PRISMA-ScR): Checklist and Explanation. Ann Intern Med 2018;169:467–73.

11 Biomarkers - MeSH - NCBI. https://www.ncbi.nlm.nih.gov/mesh/68015415 (accessed 28 Feb 2020).

12 WHO | International Classification of Diseases, 11th Revision (ICD-11). Published Online First: 11 October 2019. http://www.who.int/classifications/icd/en/ (accessed 28 Nov 2019).

13 Kivimäki M, Kuosma E, Ferrie JE, et al. Overweight, obesity, and risk of cardiometabolic multimorbidity: pooled analysis of individual-level data for 120 813 adults from 16 cohort studies from the USA and Europe. The Lancet Public Health. 2017;2:e277–85. doi:10.1016/s2468-2667(17)30074-9

14 Stirland LE, Russ TC, Ritchie CW, et al. Associations between multimorbidity and cerebrospinal fluid amyloid: a cross-sectional analysis of the European Prevention of Alzheimer’s Dementia (EPAD) V500.0 Cohort. Journal of Alzheimer’s Disease. 2019;71:703–11. doi:10.3233/jad-190222

15 Ermogenous C, Green C, Jackson T, Ferguson M, Lord JM. Treating age-related multimorbidity: the drug discovery challenge. Drug Discov Today. 2020;25(8):1403–1415. doi:10.1016/j.drudis.2020.06.016

16 Hanlon P, Hannigan L, Rodriguez-Perez J, et al. Representation of people with comorbidity and multimorbidity in clinical trials of novel drug therapies: an individual-level participant data analysis. BMC Med. 2019;17(1):201. doi:10.1186/s12916-019-1427-1

17 Buffel du Vaure C, Dechartres A, Battin C, Ravaud P, Boutron I. Exclusion of patients with concomitant chronic conditions in ongoing randomised controlled trials targeting 10 common chronic conditions and registered at ClinicalTrials.gov: a systematic review of registration details. BMJ Open. 2016;6(9):e012265. doi:10.1136/bmjopen-2016-012265

18 Lara J, Cooper R, Nissan J, et al. A proposed panel of biomarkers of healthy ageing. BMC Med. 2015;13:222. doi:10.1186/s12916-015-0470-9

19 Stirland LE, González-Saavedra L, Mullin DS, et al. Measuring multimorbidity beyond counting diseases: systematic review of community and population studies and guide to index choice. BMJ 2020;368. doi:10.1136/bmj.m160

20 Multimorbidity - MeSH - NCBI. https://www.ncbi.nlm.nih.gov/mesh/?term=multimorbidity (accessed 28 Feb 2020).

21 Aronson JK. Defining aspects of mechanisms: evidence-based mechanism (evidence for a mechanism), mechanism-based evidence (evidence from a mechanism), and mechanistic reasoning. In: LaCaze A, Osimani B (editors). Uncertainty in Pharmacology: Epistemology, Methods, and Decisions. Heidelberg Springer Verlag, 2020; Chapter 1:3–38.

## Included studies

Amaral C de A, Portela MC, Muniz PT, Farias Edos S, Araújo TS, Souza OF. Association of handgrip strength with self-reported diseases in adults in Rio Branco, Acre State, Brazil: a population-based study. Cad Saude Publica. 2015;31(6):1313–1325. doi:10.1590/0102-311X00062214

Bauml J, Chen L, Chen J, et al. Arthralgia among women taking aromatase inhibitors: is there a shared inflammatory mechanism with co-morbid fatigue and insomnia?. Breast Cancer Res. 2015;17(1):89. Published 2015 Jun 28. doi:10.1186/s13058-015-0599-7

Booth HP, Prevost AT, Gulliford MC. Impact of body mass index on prevalence of multimorbidity in primary care: cohort study. Fam Pract. 2014;31(1):38–43. doi:10.1093/fampra/cmt061

Cervellati C, Romani A, Bosi C, et al. Serum levels of hydroperoxides and multimorbidity among older patients with mild cognitive impairment or late-onset Alzheimer’s disease. Aging Clin Exp Res. 2015;27(6):799–804. doi:10.1007/s40520-015-0352-1

Cheung CL, Nguyen US, Au E, Tan KC, Kung AW. Association of handgrip strength with chronic diseases and multimorbidity: a cross-sectional study. Age (Dordr). 2013;35(3):929–941. doi:10.1007/s11357-012-9385-y

Dankel SJ, Loenneke JP, Loprinzi PD. The Impact of Overweight/Obesity Duration and Physical Activity on Medical Multimorbidity: Examining the WATCH Paradigm. Am J Health Promot. 2018;32(8):1747–1750. doi:10.1177/0890117118768893

Fabbri E, An Y, Zoli M, et al. Aging and the burden of multimorbidity: associations with inflammatory and anabolic hormonal biomarkers. J Gerontol A Biol Sci Med Sci. 2015;70(1):63–70. doi:10.1093/gerona/glu127

Fabbri E, Tanaka T, An Y, et al. Loss of Weight in Obese Older Adults: A Biomarker of Impending Expansion of Multimorbidity?. J Am Geriatr Soc. 2015;63(9):1791–1797. doi:10.1111/jgs.13608

Farup PG, Rootwelt H, Hestad K. APOE - a genetic marker of comorbidity in subjects with morbid obesity. BMC Med Genet. 2020;21(1):146. Published 2020 Jul 9. doi:10.1186/s12881-020-01082-2

Garrafa E, Casnici N, Squazzoni F, Uberti D, Marengoni A. C-reactive protein, lipoprotein (a) and cystatin C levels increase with multimorbidity in older persons. Eur J Intern Med. 2017;42:e25–e26. doi:10.1016/j.ejim.2017.04.010

Hirani V, Cumming RG, Naganathan V, et al. Associations between serum 25-hydroxyvitamin D concentrations and multiple health conditions, physical performance measures, disability, and all-cause mortality: the Concord Health and Ageing in Men Project. J Am Geriatr Soc. 2014;62(3):417–425. doi:10.1111/jgs.12693

Humphreys J, Jameson K, Cooper C, Dennison E. Early-life predictors of future multi-morbidity: results from the Hertfordshire Cohort. Age Ageing. 2018;47(3):474–478. doi:10.1093/ageing/afy005

Hyun DG, Oh YM, Lee SW, Lee SD, Lee JS. Clinical Phenotypes, Comorbidities, and Exacerbations according to Serum 25-OH Vitamin D and Plasma Fibrinogen Levels in Chronic Obstructive Pulmonary Disease. J Korean Med Sci. 2019;34(29):e195. Published 2019 Jul 29. doi:10.3346/jkms.2019.34.e195

Ishizaki T, Kobayashi E, Fukaya T, Takahashi Y, Shinkai S, Liang J. Association of physical performance and self-rated health with multimorbidity among older adults: Results from a nationwide survey in Japan. Arch Gerontol Geriatr. 2019;84:103904. doi:10.1016/j.archger.2019.103904

Issa SF, Lindegaard HM, Lorenzen T, et al. Increased serum levels of microfibrillar-associated protein 4 (MFAP4) are not associated with clinical synovitis in rheumatoid arthritis but may reflect underlying cardiovascular comorbidity. Clin Exp Rheumatol. 2020;38(1):122–128.

Kahl KG, Rudolf S, Stoeckelhuber BM, et al. Bone mineral density, markers of bone turnover, and cytokines in young women with borderline personality disorder with and without comorbid major depressive disorder. Am J Psychiatry. 2005;162(1):168–174. doi:10.1176/appi.ajp.162.1.168

Khan I, Ul-Haq Z, Taj AS, Iqbal AZ, Basharat S, Shah BH. Prevalence and Association of Obesity with Self-Reported Comorbidity: A Cross-Sectional Study of 1321 Adult Participants in Lasbela, Balochistan. Biomed Res Int. 2017;2017:1076923. doi:10.1155/2017/1076923

Kivimäki M, Kuosma E, Ferrie JE, et al. Overweight, obesity, and risk of cardiometabolic multimorbidity: pooled analysis of individual-level data for 120 813 adults from 16 cohort studies from the USA and Europe. Lancet Public Health. 2017;2(6):e277–e285. Published 2017 May 19. doi:10.1016/S2468-2667(17)30074-9

Kumar NP, Moideen K, Bhootra Y, et al. Elevated circulating levels of monocyte activation markers among tuberculosis patients with diabetes co-morbidity. Immunology. 2019;156(3):249–258. doi:10.1111/imm.13023

Lee J, Nordestgaard BG, Dahl M. Elevated ACE activity is not associated with asthma, COPD, and COPD co-morbidity. Respir Med. 2009;103(9):1286–1292. doi:10.1016/j.rmed.2009.04.003

Martin-Ruiz C, Jagger C, Kingston A, et al. Assessment of a large panel of candidate biomarkers of ageing in the Newcastle 85+ study. Mech Ageing Dev. 2011;132(10):496–502. doi:10.1016/j.mad.2011.08.001

Meems LM, de Borst MH, Postma DS, et al. Low levels of vitamin D are associated with multimorbidity: results from the LifeLines Cohort Study. Ann Med. 2015;47(6):474–481. doi:10.3109/07853890.2015.1073347

Mendes A, Tezenas du Montcel S, Levy M, et al. Multimorbidity Is Associated with Preclinical Alzheimer’s Disease Neuroimaging Biomarkers. Dement Geriatr Cogn Disord. 2018;45(5-6):272–281. doi:10.1159/000489007

Moo IH, Kam CJW, Cher EWL, et al. The effect of the comorbidity burden on vitamin D levels in geriatric hip fracture. BMC Musculoskelet Disord. 2020;21(1):524. Published 2020 Aug 8. doi:10.1186/s12891-020-03554-1

Mounce LTA, Campbell JL, Henley WE, Tejerina Arreal MC, Porter I, Valderas JM. Predicting Incident Multimorbidity. Ann Fam Med. 2018;16(4):322–329. doi:10.1370/afm.2271

Niedzwiedz CL, Katikireddi SV, Pell JP, Smith DJ. Sex differences in the association between salivary telomere length and multimorbidity within the US Health & Retirement Study. Age Ageing. 2019;48(5):703–710. doi:10.1093/ageing/afz071

Pérez LM, Hooshmand B, Mangialasche F, et al. Glutathione Serum Levels and Rate of Multimorbidity Development in Older Adults [published online ahead of print, 2019 Apr 25]. J Gerontol A Biol Sci Med Sci. 2019;glz101. doi:10.1093/gerona/glz101

Sarkar C, Dodhia H, Crompton J, et al. Hypertension: a cross-sectional study of the role of multimorbidity in blood pressure control. BMC Fam Pract. 2015;16:98. Published 2015 Aug 7. doi:10.1186/s12875-015-0313-y

Schöttker B, Saum KU, Jansen EH, Holleczek B, Brenner H. Associations of metabolic, inflammatory and oxidative stress markers with total morbidity and multi-morbidity in a large cohort of older German adults. Age Ageing. 2016;45(1):127–135. doi:10.1093/ageing/afv159

Stirland LE, Russ TC, Ritchie CW, Muniz-Terrera G; EPAD Consortium. Associations Between Multimorbidity and Cerebrospinal Fluid Amyloid: A Cross-Sectional Analysis of the European Prevention of Alzheimer’s Dementia (EPAD) V500.0 Cohort. J Alzheimers Dis. 2019;71(2):703–711. doi:10.3233/JAD-190222

Strandberg AY, Strandberg TE, Stenholm S, Salomaa VV, Pitkälä KH, Tilvis RS. Low midlife blood pressure, survival, comorbidity, and health-related quality of life in old age: the Helsinki Businessmen Study. J Hypertens. 2014;32(9):1797–1804. doi:10.1097/HJH.0000000000000265

Townsend MH, Bologna NB, Barbee JG. Heart rate and blood pressure in panic disorder, major depression, and comorbid panic disorder with major depression. Psychiatry Res. 1998;79(2):187–190. doi:10.1016/s0165-1781(98)00036-5

Wong MC, Wang HH, Cheung CS, et al. Factors associated with multimorbidity and its link with poor blood pressure control among 223,286 hypertensive patients. Int J Cardiol. 2014;177(1):202–208. doi:10.1016/j.ijcard.2014.09.021

Vassilaki M, Aakre JA, Mielke MM, et al. Multimorbidity and neuroimaging biomarkers among cognitively normal persons. Neurology. 2016;86(22):2077–2084. doi:10.1212/WNL.0000000000002624

Xu X, Mishra GD, Dobson AJ, Jones M. Short-term weight gain is associated with accumulation of multimorbidity in mid-aged women: a 20-year cohort study. Int J Obes (Lond). 2019;43(9):1811–1821. doi:10.1038/s41366-018-0250-7

