## Supplementary material for "Biomarkers in the prediction of multimorbidity: scoping review": Table 2. Characteristics of included studies.

Table 2. Characteristics of included studies, by biomarker group.

| Study ID (author and year) | Prospective or cross-sectional data reported | Country and setting | Population | Sample size | Age of population at study baseline | Sex distribution of study population | Biomarker assessed | Biomarker measurement methods | Duration of follow-up | Outcome (description of multimorbidity investigated) | Ascertainment method for outcomes | Results |
|---|---|---|---|---|---|---|---|---|---|---|---|---|
| **(i) Serum biomarkers** | | | | | | | | | | | | |
| Bauml 2015 | cross-sectional | cohort of breast cancer patients, USA | adults with early-stage breast cancer | 203 | mean 60.5 years | 100% female | inflamatory biomakers: C-reactive protein, eotaxin, monocyte chemoattractant protein, vitamin D–binding protein. | multiplex assay | N/A | composite variable of arthralgia, insomnia and fatigue vs none of these; study not focused on MM, but looks at comorbidities of breast cancer among those taking aromatase inhibitors | self report using pain inventory and insomnia severity index | Multivariate analysis showed strong assoc. of inflammatory biomarkers with MM: CRP (β = 93.1; 95%CI: 25.1 to 161.1; p=0.008), eotaxin (β = 79.9; 95 % CI: 32.5 to 127.2; p = 0.001), monocyte chemoattractant protein (β =151.2; 95% CI: 32.7 to 269.8; p=0.013), and vitamin D–binding protein (β = 19,422; 95 % CI: 5500.5 to 33,344; p=0. 006) |
| Cervellati 2015 | cross-sectional | medical centre based study, Italy | outpatients referring to the Day Service for Cognitive Decline, Ferrara, Italy | 225 | age 60 upwards, approx mean 75 years | approx 60% female | serum levels of hydroperoxides | fasting venous blood samples; colorimetric assay based on the reaction between these lipid peroxidation by-products and N,N-diethyl-para-phenylenediamine | N/A | cumulative illness rating scale- comorbidity index (CIRS-CI); 14 items: heart, vascular, hematopoietic, respiratory; eyes, ears, nose; throat and larynx; upper gastrointestinal, lower gastrointestinal, liver, renal, genitourinary, musculoskeletal, neurological, endocrine-metabolic, and behavioral-psychiatric. Each condition scored: 0, no problem; 1, mild problem; 2, moderate disability or morbidity; 3, severe/constant significant disability/chronic problems; 4, life-threatening problems. | structured interview with patients and caregivers | The peroxidation marker OxS was positively & significantly correlated with multimorbidity (CIRS-CI) in controls (p = 0.002) and in mild cognitive impairment group (p = 0.005) but not in late onset Alzheimer's disease patients (p = 0.104). |
| Fabbri 2015a | prospective & cross-sectional | population based, Chianti, Italy | random selection of population aged 60+ years invited to participate | 1,018 | 73.6 ± 7.2 years | 57.2% female | inflammatory and hormonal biomarkers IL-6, IL-1ra, TNF-α receptor II (TNFAR2), and dehydroepiandrost erone sulfate | High-sensitivity ELISA, multiplex, chemiluminescent immunoassays. | 9 years | 2 or more of: hypertension, diabetes mellitus, coronary artery disease, congestive heart failure, stroke, COPD, cancer, Parkinson's disease, hip fracture, lower extremity joint disease, anaemia, renal failure, peripheral arterial disease, cognitive impairment, depression | self-report at study visit | Cross-sectional findings: higher IL-6, IL-1ra, TNF-α receptor II (TNFAR2), and lower DHEA were associated with higher number of diseases, independent of age, sex, body mass index, and education. Prospective findings: higher baseline IL-6 and steeper increase of IL-6 levels were significantly and independently associated with a steeper increase in MM over time (p <001 and p = .003, respectively) |

Table 2. Characteristics of included studies, by biomarker group.

| | | | | | | | | | | | | |
|---|---|---|---|---|---|---|---|---|---|---|---|---|
| Garrafa 2017 | cross-sectional | prospective study, Italy | community elderly | 134 | mean 77.7 (SD 7.6) years | 59.3% female | serum C-reactive protein, lipoprotein (a) & cystatin C levels | latex lipoprotein reagent; immunoassay Cyst-C nd CRP | N/A | no. chronic diseases | NR | Higher value of all the three biomarkers correlated with an increase in the number of chronic conditions, both when dichotomized as "high" versus normal and as continuous variables. |
| Hirani 2014 | cross-sectional | population study, Sydney, Australia | community dwelling men aged 70+ years | 1,659 | mean 77, range 70 to 97 | 100% male | serum 25-hydroxyvitamin D | manual radio-immunoassay | N/A | High comorbid burden was defined as the presence of four or more of: diabetes mellitus, thyroid dysfunction, osteoporosis, Paget's disease, stroke, Parkinson's disease, epilepsy, hypertension, heart attack, angina pectoris, congestive heart failure, intermittent claudication, chronic obstructive lung disease, liver disease, cancer (excluding nonmelanoma skin cancers), osteoarthritis, or gout. | self-report of diagnosis | Odds of having 4+ conditions vs 0 to 3 associated with low serum vit D: <50.0 nmol/L vs ref ≥75 nmol/L 25(OH)D OR=1. 52 (95%CI 1.11 to 2.10), p=0. 01; 50.0 to 74.9 nmol/L vs ref ≥75 nmol/L 25(OH)D OR 1.11 (95%CI 0.80 to 1.55), p=0.52 |
| Hyun 2019 | cross-sectional | oupatients at respiratory clinic, Seoul, S Korea | outpatients with COPD | 236 | approx 69 years | approx 10% female | plasma fibrinogen and serum 25-OH vitamin D | plasma fibrinogen concentrations were determined via the Clauss assay; 25-OH vitamin D levels were measured via radioimmuno-assay | N/A | diabetes mellitus in addition to COPD | electronic health record | Among people with COPD, patients with high plasma fibrinogen concentrations and normal 25-OH vitamin D levels had a significantly higher incidence of DM than did the other patients. |
| Issa 2020 | prospective | outpatient setting, Denmark | Patients with newly diagnosed or longstanding rheumatoid arthritis | 364 (317 long-standing, 47 newly diagnosed) | 18 to 71 years newly diagnosed; 19 to 84 years long-standing RA | 62% women | MFAP4 | serum and synovial fluid MFAP4 was measured using an AlphaLISA immunoassay | 4 years | Cardiovascular comorbidity (among rheumatoid arthritis sufferers) | Cardiovascular co-morbidity details were extracted from relevant databases in the National Patient Registry and from hospital databases. | MFAP4 correlated positively with stroke events (p=0.08), systolic blood pressure (p<0. 001) and levels of HDL cholesterol (p<0.01). In multivariate analysis, adjusting for age, gender and smoking, only systolic blood pressure remained significantly associated with MFAP4, p=0. 001 |

Table 2. Characteristics of included studies, by biomarker group.

| | | | | | | | | | | | | |
|---|---|---|---|---|---|---|---|---|---|---|---|---|
| Kahl 2005 | cross-sectional | Inpatient psychiatric unit, Germany | patients with borderline personality disorder and controls | 58 | approx mean 26 years | 100% female | bone mineral density, also markers of bone turnover, and endocrine and immune measures crosslaps, osteocalcin, serum cortisol, tumor necrosis factor-α (TNF-α), and interleukin-6 | Bone mineral density was measured in all patients by means of dual-energy x-ray absorptiometry at the lumbar spine, right femur, left femur, and the forearm of the nondominant hand | N/A | borderline personality disorder with major depressive disorder (vs borderline personality disorder alone) | Structured Clinical Interview for DSM-IV (SCID) and the SCID for Personality Disorders. | BMD significantly lower in group with borderline personality disorder and comorbid major depressive disorder than in group with borderline personality disorder alone. Values of crosslaps, osteocalcin, serum cortisol, TNF-α, and interleukin-6 were significantly higher in the patients with borderline disorder plus current major depressive episode than in the healthy subjects; Patients with borderline personality disorder who did not have current or lifetime depression displayed no alterations of either bone mineral density or the immunological and hormonal measures examined. |
| Kumar 2019 | cross-sectional | Chennai, India | 44 ppts with active pulmonary TB with DM, 44 ppts with active pulmonary TB, 44 ppts with diabetes mellitus, and 30 healthy control ppts with no TB or diabetes | 132 | approx mean 50 years, range 22 to 70 years | approx 40% female | monocyte activation markers | Circulating levels of monocyte subsets by ELISA: sCD14, sCD163, CRP and sTF | N/A | TB with concurrent diabetes mellitus | Smear and culture positivity for M. tuberculosis. Chest cavity X ray | TB-with diabetes was associated with elevated systemic levels of circulating monocyte activation markers compared with TB alone. |
| Martin-Ruiz 20 | cross-sectional | Newcastle, UK | cross sectional analysis of ongoing prospective study | 852 | 85 years | 61% female | 74 candidate biomarkers | anthropometry, spirometry, sphygmomanometry, blood and serum samples analysed for inflammatory, immune, nutrition markers and DNA markers | N/A | disease count from 18 possible: hypertension, ischaemic heart disease, cerebrovascular disease, peripheral vascular disease, heart failure, atrial flutter or fibrillation, arthritis, osteoporosis, chronic obstructive pulmonary disease or asthma, other respiratory disease, diabetes, hypothyroidism or hyperthyroidism, cancer diagnosed in past five years (excluding non-melanoma skin cancer), eye disease, dementia, [illegible] | research nurse interview and general practice electronic records | Many biomarkers associated with disease; the most strongly associated with multimorbidity were: BNP (N-terminal proB-type natriuretic peptide), handgrip strength, bp, timed up-and-go, FEV, haematocrit, haemoglobin, rbc count, free T3, vitamin D. |

Table 2. Characteristics of included studies, by biomarker group.

| | | | | | | | | | | | | |
|---|---|---|---|---|---|---|---|---|---|---|---|---|
| Meems 2015 | cross-sectional | prospective cohort, The Netherlands | men & women aged 25 to 50, and older and younger family members | 8,726 | 45 =/-13 years, | 73% female | plasma 25-hydroxyvitamin D3 | Plasma 25-hydroxyvitamin D3 levels were measured by solid phase extraction isotope dilution followed by liquid chromatography – tandem mass spectrometry | N/A | 2 or more conditions within disease domains: genitourinary, renal, hematologic, dermatologic, musculoskeletal, ophthalmic, ENT, psychiatric, endocrine, cardiovascular, respiratory, CNS, gastrointestinal disease. | self-report plus clinical examination | Each incremental reduction by 1 SD of vitamin D level was associated with an 8% higher morbidity score (full model OR 0.92, 95%CI 0.88 to 0.97, P =0.001). Participants with vitamin D levels 25 nmol/L were at highest risk for increasing morbidity prevalence (versus 80 nmol/L, OR 1.34, 95%CI 1.07 to 1.67, P=0.01). |
| Moo 2020 | cross-sectional | hospital, Singapore | Patients with hip fracture admitted to hospital: age > 60 years with femoral neck, intertrochanteric, or subtrochanteric fractures after a low-energy fall. | 796 | mean 77.7 ± 8.0 years | 71% women | serum 25-hydroxyvitamin D3 | radioimmunoassay method, as part of the institution hip fracture protocol | N/A | Charlson Comorbidity Index | hospital records | Mean vit D level was 20.4 ± 7.4 ng/mL. There was no correlation between serum vit D level and age-adjusted Charlson Comorbidity Index. The 8% of patients with low comorbidity burden had a mean 25(OH)D level of 20.1 ± 7.2 ng/mL; the remaining 92% of the patients with high comorbidity burden had a mean 25(OH)D level of 20.5 ± 7.4. |
| Perez 2019 | prospective | population based, Sweden | ≥60-year-olds living in Stockholm | 2,596 | mean 82.8 (SD 10.2) years | 61.3% female | serum glutathione | nonfasting venous bloods taken; levels of total serum glutathione (µmol/L), including both the reduced and oxidized form of glutathione, were measured through tandem mass spectroscopy. | 6 years | no. chronic conditions in 60 broad disease categories | Nurses collected demographic data & assessed physical function; psychologists administered cognitive test batteries; physicians carried out physical, geri-atric, and neuropsychiatric examinations. Data on past medical his-tory and vital status from National Patient Register & Swedish Death Register. | Lower levels of baseline total serum glutathione were associated with a higher rate of multimorbidity development. The direction and magnitude of the association remained very stable regardless of potential confounders |

Table 2. Characteristics of included studies, by biomarker group.

| | | | | | | | | | | | | |
|---|---|---|---|---|---|---|---|---|---|---|---|---|
| Schöttker 2016 | cross-sectional | population based prospective study cohort, Germany | mid-age to eldery adults | 2,547 | 70 (SD 6) years | 50% female | metabolic, inflammatory and oxidative stress markers | full blood HbA1c & serum C-reactive protein by immunoturbidimetry. Serum total cholesterol, HDL-cholesterol & triglycerides, by enzymatic chromatography. Derivatives of reactive oxygen metabolites (proxy for ROS production) and total thiol levels (proxy for the redox status of blood), specific assays. | N/A | multi-morbidity = two or more of 13 somatic organ systems. | Total somatic morbidity & multi-morbidity from GP questionnaire with the Cumulative Illness Rating Scale-Geriatric version. | All markers except total thiol were significantly associated with multi-morbidity; suggested U-shaped associations. |
| **(ii) Molecular biomarkers** | | | | | | | | | | | | |
| Lee 2009 | prospective | general population, Denmark | men and women | 9,034 | age 20+ years | 55% female | ACE polymorphisms | Insertion/deletion polymorphism of 287 bp in intron 16 of the ACE gene was identified by conventional polymerase chain reaction (PCR) using two primers flanking the site of the insertion. All samples apparently homozygous for the D allele were subjected to a second PCR amplification with an insertion-specific primer. | NR | COPD + asthma/IHD/hypertension vs COPD alone | self report of asthma, spirometry for COPD, ischaemic heart disease diagnoses from Danish National Hospital Discharge Registry, blood pressure by sphygmomanometry | No evidence of association between ACE I/D genotype and MM: OR for (IHD + COPD) vs COPD alone by genotype: 1.1 (0.8 to 1.6) for ID and 1.2 (0.8 to 0.7) for DD compared with II individuals; OR for (hypertension + COPD) vs COPD alone: 1.1 (0.7 to 1.5) for ID and 0.8 (0.5 to 1.2) for DD compared with II individuals; OR for (asthma + COPD) vs COPD alone: 1.2 (0.9 to 1.4) for ID and 1.2 (0.9 to 1.5) for DD compared with II individuals. |
| Niedzwiedz 2019 | prospective & cross-sectional | US community survey | individuals aged 50+ years in the US | 5,495 | mean age men = 66.8 years, women = 67.6 years | 52% female | salivary telomere length | quantitative Polymerase Chain Reaction (qPCR) which compared the telomere sequence copy number in each individual's sample (T) to a single-copy gene copy number (S), producing a T/S ratio which is proportional to telomere length. | 2 to 6 years | 2 or more of: high blood pressure; diabetes or high blood sugar; cancer or a malignant tumour (excluding minor skin cancer); chronic lung disease (e.g. chronic bronchitis or emphysema); heart attack, coronary heart disease, angina, congestive heart failure, or other heart problems; stroke; emotional, nervous, or psychiatric problems; arthritis or rheumatism. | self report of doctor's diagnosis | In the cross-sectional logistic regression analyses, telomere length was related to reduced likelihood of multimorbidity in men (OR=0.88, 95% CI: 0.63 to 1.25) and women (OR=0.97, 95% CI: 0.72 to 1.31), but these associations were not statistically significant when using the binary multimorbidity variable as the outcome. |

Table 2. Characteristics of included studies, by biomarker group.

| | | | | | | | | | | | | |
|---|---|---|---|---|---|---|---|---|---|---|---|---|
| **(iii) Physiological biomarkers** | | | | | | | | | | | | |
| Amaral 2015 | cross-sectional | adult population survey, Acre State, Brazil | random selection of adult population | 1,395 | 18 to 96 years | 54.6% female | handgrip strength | dynamometry: higher of 2 attempts with dominant hand | N/A | 2 or more of: hypertension, diabetes mellitus, MI/stroke, dyslipidemia, depression, chronic kidney disease, musculoskeletal disorders | self-report of physician diagnosis | among men, multimorbidity OR for low versus high handgrip strength = 1.99 (1.27 to 3.12); among women the association was not significant |
| Cheung 2013 | cross-sectional | Hong Kong Osteoporosis Study | community aged 50+ years | 1,145 | 50+ years | 35% female | handgrip strength | dynamometry: mean of 3 attempts | N/A | 2 or more of 18 conditions: anaemia, anxiety, cataract, stroke, CKD stage 3 or above,, COPD, depression, diabetes, history of fall in past 12 months, hepatitis B, hyperlipidaemia, hypertension, hyperthyroidism, ischemic heart diseases, kyphosis, malignancy within 5 years, osteoarthritis knee, peptic ulcer. | Questionnaire administered by study staff; medical records confirmed by electronic health record | Handgrip strength was associated with multiple chronic diseases and multimorbidity in men and women, after adjustment for confounding factors.There was a linear trend of association with the number of chronic diseases in men, but not in women. |
| Ishizaki 2019 | cross-sectional | Japan, nationwide longitudinal survey | adults aged ≥60 years | 2,525 | 60+ years | 54% female | handgrip strength | grip strength (measured in the standing position twice for each hand using a Smedley hand dynamometer) and walking speed (measured thrice over 2.5m) assessed during a home visit by study staff | N/A | 2 or more of: heart disease, arthralgia, hypertension, diabetes, stroke, cataract, cancer, respiratory disease, and low back pain | Self-report in ongoing longitudinal study | Adjusting for demographic and lifestyle variables, multimorbidity was significantly associated with poor grip strength (P=0.006) but not with walking speed (P=0.479). |
| Sarkar 2015 | cross-sectional | patients with hypertension , primary care, UK | Lambeth Data-Net, a patient level primary care database | 31,676 | mean 63.9 (SD 14.2) years | 53.8% female | bp | electronic primary care record | N/A | ischaemic heart disease, heart failure, diabetes mellitus, chronic kidney disease, stroke and atrial fibrillation; depression, serious mental illness, dementia; chronic obstructive pulmonary disease, asthma; epilepsy. | electronic primary care record | MM was associated with lower bp: in participants with one morbidity [in addition to hypertension], mean bp was 137.1 mmHg (95%CI 136.7 to 137.4); bp among those with MM (i.e. two comorbidities) was 136.0 mmHg (95 % CI 135.5 to 136.5) and for those with three comorbidities was 134.3 mmHg (95 % CI 133.5 to 135.2). |
| Strandberg 2014 | prospective | Finland, population based | business executives | 3,267 | mean 40 years | 100% male | blood pressure | sphygmo-manometry | 48 years follow-up | diseases including diabetes and hypertension | self-reported | Baseline BP had a graded and highly significant association with number of comorbidities (P < 0.001). |

Table 2. Characteristics of included studies, by biomarker group.

| | | | | | | | | | | | | |
|---|---|---|---|---|---|---|---|---|---|---|---|---|
| Townsend 199 | cross-sectional | patients with panic disorder, New Orleans, USA | patients with panic disorder recruited to join study | 76 | 38.2 (SD 9.75) years | 63.6% female | heart rate variability and bp | bp and heart rate using Omron digital blood pressure monitor | N/A | panic disorder with or without major depression | NR | BP and heart rate higher in MM group (panic disorder + major depression) compared with panic disorder alone or compared with major depression alone |
| Wong 2014 | cross-sectional | Hong Kong | patients with hypertension | 223,286 | 59.9 (SD 17.6) years | 54.8% female | bp | electronic record of hypertensive medication, updated with study clinic measurements by sphygmanometry | N/A | multimorbidity cardiovascular, respiratory, diabetes/impaired fasting glucose, renal disease | electronic health record | Patients with poor blood pressure control far more likely to have one (65.0% vs 35.0%, AdjOR 3.38, 95%CI 3.29 to 3.46, p <0.001) or ≥2 comorbities (69.1% vs 30.9%, AdjOR 4.49, 95%CI 4.28 to 4.70, p < 0.001) concomitant medical conditions. |
| **(iv) Body size biomarkers** | | | | | | | | | | | | |
| Booth 2014 | prospective | UK GP databases (CPRD) | In CPRD, a random sample of men and women aged ≥30 years from the years 2005–11. | 223,089 | ≥30 years | NR | BMI | electronic health record - CPRD | 1 to 6 years | two or more concurrent conditions from: coronary heart disease, stroke, asthma, sleep apnoea, type 2 diabetes, all neoplasms, gallbladder, back pain, osteoarthritis, other joint problems, depression. | electronic health record - CPRD | Multimorbidity was highly associated with increasing BMI category and obesity. In a cross-sectional analysis, 32% of multimorbidity was attributable to overweight and obesity. |
| Dankel 2018 | cross-sectional | national health survey, USA | adults across the US | 3,621 | 36 to 85 yrs | NR | BMI | height and weight measured by study staff | N/A | 2 or more of: arthritis, asthma, bronchitis, cancer, congestive heart failure, coronary artery disease, diabetes, emphysema, liver disease, stroke, high total cholesterol (>240 mg/dL), low high density lipoprotein levels (<40 mg/dL), and hypertension (140/90 mm Hg). | self-report of physician diagnosis | The odds of MM were increased as the classification and duration of overweight/obesity worsened |
| Fabbri 2015b | prospective & cross-sectional | population based, Chianti, Italy | random selection of population invited to participate | 1,025 | 73.8 (± 7.2) years | 55.8% female | BMI | study staff measured height and weight at study visit | mean 4 years | 2 or more of: hypertension, diabetes mellitus, coronary artery disease, congestive heart failure, stroke, COPD, cancer, Parkinson's disease, hip fracture, lower extremity joint disease, anaemia, renal failure, peripheral arterial disease, cognitive impairment, depression | self-report at study visit | Higher baseline BMI was associated with greater longitudinal increase in number of chronic diseases. Greater decline in BMI tended to be associated with increase in multimorbidity, independent of baseline BMI. Baseline obesity was associated with high baseline MM. After adjusting for age and sex, obese participants had significantly more diseases (1.9) than nonobese participants (1.6). No significant difference in number of diseases was found between overweight and normal-weight participants. |

Table 2. Characteristics of included studies, by biomarker group.

| | | | | | | | | | | | | |
|---|---|---|---|---|---|---|---|---|---|---|---|---|
| Humphreys 2018 | prospective | UK, community | middle aged men & women | 2091 | mean 66 years | NR | BMI | NR | mean 7 years | total no. of multimorbid conditions vs no MM. Data on adult BMI extracted though focus of paper is on early life exposures. | self report | Higher BMI by 1kg/m2 associated with increased risk MM: OR 1.12 (95% CI 1.10-1.15) |
| Khan 2017 | cross-sectional | population based, Balochistan, Pakistan | stratified random sampling of adult population | 1,321 | mean 32.2 (SD 12.8) yrs | 50.1% female | BMI, waist circumference, waist to hip ratio | study staff anthropometry, standard operating procedures, mean of three measures | N/A | 2 or more of: coronary heart disease or stroke, hypertension, hypercholesterolemia, type 2 DM, osteoarthritis, cancer | self-report of a doctor's diagnosis or taking medication prescribed by a doctor for | Significant linear trend of increasing morbidities with increasing body weight categories. MM not the focus of this paper. |
| Kivimaki 2017 | prospective | USA and Europe | various populations, community based | 120,813 | mean 51.4 years | 40%M 60% F | BMI | measured or self reported | mean 10.7 years | incident cardiometabolic MM (i.e. CHD, stroke, diabetes) | self report or electronic medical records | For overweight, obesity and severe obesity (BMI 35+kg/m2) compared with normal weight, OR =2.0 (95% CI 1.7-2.4); 4.5 (3.5-5.8) and 14.5 (10.1-21.0). |
| Mounce 2018 | prospective | England, UK, community | middle aged men & women | 1,477 | 50+ years | NR | BMI | nurse measured | 6-7 years | HR of MM compared with no morbidity | self report | Obesity associated with increased risk of incident MM: obese vs normal weight HR=1.92 (95% CI 1.43 - 2.59). Data on BMI extracted; paper reports a range of exposures |
| Xu 2019 | prospective | Australian community | mid-age women living in Australia | 7,357 | mean 4 years | 100% female | BMI change | self-reported height and weight | 20 years | 2 or more of: nine conditions: cancer, cardiovascular disease, hypertension, depression/anxiety, diabetes mellitus, asthma, chronic obstructive pulmonary disease, arthritis, osteoporosis. | self-report | Baseline BMI, time-varying BMI and short-term weight change were all associated with the accumulation of multimorbidity. High weight gain was associated with a 25% increased odds of multimorbidity: OR 1.25 (95% CI 1.08 to 1.45) compared with maintaining a stable weight. |
| **(v) Brain function markers** | | | | | | | | | | | | |

Table 2. Characteristics of included studies, by biomarker group.

| | | | | | | | | | | | | |
|---|---|---|---|---|---|---|---|---|---|---|---|---|
| Mendes 2018 | cross-sectional | prospective cohort, France | men and women aged 70 to 85 years at baseline | 318 | mean 76 (SD 3) years | 64.2% female | neuropsychological and neuroimaging assessment with automated methods that measured hippocampal volumes, white matter hyperintensity volumes, fluorodeoxyglucose positron emission tomography (FDG-PET) standardized uptake values (SUV) in AD signature regions, and amyloid positron emission tomography (amyloid-PET) SUV ratios | MRI and PET scan of brain | N/A | 2 or more of: hypertension, dyslipidemia, diabetes mellitus, atrial fibrillation, heart failure, chronic kidney disease, obstructive sleep apnea, active or past smoking, unhealthy alcohol consumption, prior head trauma, obesity, vitamin B12 deficiency, depression and post-traumatic stress disorder. | self report of diagnosis; validated by physician | Accumulation of multimorbidity, was associated with neuroimaging markers of AD neurodegeneration, but not with amyloid deposition. Increasing no. comorbidities was sig. associated with lower hippocampal volumes (-0.03 ±0.01; P = .012; R2 = .017) as well as with lower SUV (-0.027 ±0.009; P = .005; R2 = .022) in FDG-PET. There was no assoc. between comorbidities accumulation & SUVr in amyloid PET (0.001 ±0.007; P =0.884; R2 = .0001). After adjustment for possible confounding factors, the assoc. remained statistically sig. only for FDG-PET SUV (-0.02 ±0. 01; P = .038; R2 = .07). |
| Stirland 2019 | cross-sectional | Europe, prospective cohort | volunteers aged 50+ at 12 sites across Europe | 447 | 66.6 (SD 6.6) yrs | 52.3% women | cerebrospinal fluid amyloid-β42 | lumbar puncture samples assayed by immunoassay | N/A | 2 or more of 39 chronic conditions | doctor assessed medical history | People with MM were approx. 40% less likely to be amyloid positive than people with 0–1 conditions (OR adjusted for age, sex, APOE status, family history of dementia and years of education 0.59, 95%CI 0.37–0.95, p = 0.030). Compared to participants with 0–1 conditions, those with MM had an increase in Abeta42 by 115.7 pg/ml, but this was not statistically significant (95% CI –7.7 to 239.0, p = 0.066). |
| Vassilaki 2016 | cross-sectional | cohort study, Minnesota, USA | Minnesota residents aged 70 to 89 at baseline with no cognitive impairment | 1,449 | mean 79 yrs | 40.1% female | imaging biomarkers of brain pathology | MRI and PET scan of brain | N/A | 2 or more of: hyperlipidemia, diabetes mellitus, hypertension, cardiac arrhythmias, coronary artery disease, stroke, congestive heart failure, cancer, asthma, depression, substance abuse disorders (drugs and alcohol), COPD, chronic kidney disease, arthritis, osteoporosis, schizophrenia, and hepatitis. | electronic health record | Multimorbidity and severe multimorbidity (4+ chronic conditions) were associated with abnormal Alzheimer disease (AD) signature meta–region of interest (meta-ROI) 18F-FDG hypometabolism (OR 2.03; 95% CI 1.10 to 3.77 and OR 2.22; 95% CI 1.18 to 4.16, respectively), and with abnormal AD signature MRI cortical thickness (OR 1.53; 95% CI 1.09 to 2.16 and OR 1.76; 95% CI 1.24 to 2.51, respectively), but was not associated with amyloid accumulation. |
